# Whole-Genome Sequencing Analysis Reveals New Susceptibility Loci and Structural Variants Associated with Progressive Supranuclear Palsy

**DOI:** 10.1101/2023.12.28.23300612

**Authors:** Hui Wang, Timothy S Chang, Beth A Dombroski, Po-Liang Cheng, Vishakha Patil, Leopoldo Valiente-Banuet, Kurt Farrell, Catriona Mclean, Laura Molina-Porcel, Alex Rajput, Peter Paul De Deyn, Nathalie Le Bastard, Marla Gearing, Laura Donker Kaat, John C Van Swieten, Elise Dopper, Bernardino F Ghetti, Kathy L Newell, Claire Troakes, Justo G de Yébenes, Alberto Rábano-Gutierrez, Tina Meller, Wolfgang H Oertel, Gesine Respondek, Maria Stamelou, Thomas Arzberger, Sigrun Roeber, Ulrich Müller, Franziska Hopfner, Pau Pastor, Alexis Brice, Alexandra Durr, Isabelle Le Ber, Thomas G Beach, Geidy E Serrano, Lili-Naz Hazrati, Irene Litvan, Rosa Rademakers, Owen A Ross, Douglas Galasko, Adam L Boxer, Bruce L Miller, Willian W Seeley, Vivanna M Van Deerlin, Edward B Lee, Charles L White, Huw Morris, Rohan de Silva, John F Crary, Alison M Goate, Jeffrey S Friedman, Yuk Yee Leung, Giovanni Coppola, Adam C Naj, Li-San Wang, PSP genetics study group, Dennis W Dickson, Günter U Höglinger, Gerard D Schellenberg, Daniel H Geschwind, Wan-Ping Lee

## Abstract

**Background:** Progressive supranuclear palsy (PSP) is a rare neurodegenerative disease characterized by the accumulation of aggregated tau proteins in astrocytes, neurons, and oligodendrocytes. Previous genome-wide association studies for PSP were based on genotype array, therefore, were inadequate for the analysis of rare variants as well as larger mutations, such as small insertions/deletions (indels) and structural variants (SVs).

**Method:** In this study, we performed whole genome sequencing (WGS) and conducted association analysis for single nucleotide variants (SNVs), indels, and SVs, in a cohort of 1,718 cases and 2,944 controls of European ancestry. Of the 1,718 PSP individuals, 1,441 were autopsy-confirmed and 277 were clinically diagnosed.

**Results:** Our analysis of common SNVs and indels confirmed known genetic loci at *MAPT*, *MOBP*, S*TX6*, *SLCO1A2*, *DUSP10*, and *SP1*, and further uncovered novel signals in *APOE*, *FCHO1/MAP1S, KIF13A, TRIM24, TNXB, and ELOVL1*. Notably, in contrast to Alzheimer’s disease (AD), we observed the *APOE* ε2 allele to be the risk allele in PSP. Analysis of rare SNVs and indels identified significant association in *ZNF592* and further gene network analysis identified a module of neuronal genes dysregulated in PSP. Moreover, seven common SVs associated with PSP were observed in the H1/H2 haplotype region (17q21.31) and other loci, including *IGH*, *PCMT1*, *CYP2A13*, and *SMCP*. In the H1/H2 haplotype region, there is a burden of rare deletions and duplications (*P* = 6.73×10^-3^) in PSP.

**Conclusions:** Through WGS, we significantly enhanced our understanding of the genetic basis of PSP, providing new targets for exploring disease mechanisms and therapeutic interventions.

## Background

Progressive supranuclear palsy (PSP) is a neurodegenerative disease that is pathologically defined by the accumulation of aggregated tau protein in multiple cortical and subcortical regions, especially involving the basal ganglia, dentate nucleus of the cerebellum midbrain [1]. An isoform of tau harboring 4 repeats of microtubule-binding domain (4R-tau) is particularly prominent in these tau aggregates [2]. Clinical manifestations of PSP include a range of phenotypes, including the initially described and most common, PSP-Richardson syndrome that presents with multiple features, including postural instability, vertical supranuclear palsy, and frontal dementia. However, there are several other phenotypes, such as PSP-Parkinsonism, PSP-Frontotemporal dementia, PSP-freezing of gait, PSP-speech and language disturbances, etc. [3]. Presentation of these phenotypes varies widely depending on the distribution and severity of the pathology [4–6].

Currently, the most recognized genetic risk locus for PSP is at the H1/H2 haplotype region covering *MAPT* gene at chromosome 17q21.31 [7], where individuals carrying the common H1 haplotype are more likely to develop PSP with an estimated odds ratio (OR) of 5.6 [8]. Previous studies usually ascribed the observed association in the H1/H2 haplotype to *MAPT* [7,9,10]. However, recent functional dissection of this region using multiple parallel reporter assays coupled to CRISPRi demonstrated multiple risk genes in the area in addition to *MAPT*, including *KANSL1* and *PLEKMHL1* [11]. Genome-wide association studies (GWASs) in PSP have identified common variants in *STX6*, *EIF2AK3*, *MOBP*, *SLCO1A2*, *DUSP10*, *RUNX2*, and *LRRK2* with moderate effect size [8,12–14]. In addition, variants in *TRIM11* were identified as a genetic modifier of the PSP phenotype when comparing PSP with Richardson syndrome to PSP without Richardson syndrome [15].

To date, no comprehensive analysis of single nucleotide variants (SNVs), small insertions and deletions (indels), and structural variants (SVs) in PSP by whole genome sequencing has been conducted. To gain a more comprehensive understanding of the genetic underpinnings of PSP, we performed whole genome sequencing (WGS) and analyzed SNVs, indels and SVs. As a result, we not only validated previously reported genes but also unveiled new loci that provide novel insights into the genetic basis of PSP.

## Methods

### Study subjects

We performed WGS at 30x coverage (**Table S1**) for 1,834 PSP cases and 128 controls from the PSP-NIH-CurePSP-Tau, PSP-CurePSP-Tau, PSP-UCLA, and AMPAD-MAYO cohorts included in Alzheimer’s Disease Sequencing Project (ADSP, NG00067.v7) and used 3,008 controls from other cohorts in ADSP [16]. Control subjects were self-identified as non-Hispanic white. WGS data is available on The National Institute on Aging Genetics of Alzheimer’s Disease Data Storage Site (NIAGADS) [17]. We removed related subjects (identify by descent > 0.25), five clinically diagnosed PSP who were not found to have PSP on autopsy, and non-Europeans (subjects that were eight standard deviations away from the 1000 Genomes Project European samples [18,19] using the first six principal components (PCs)), resulting in 1,718 individuals with PSP and 2,944 control subjects. Of the 1,718 PSP individuals, 1,441 were autopsy-confirmed and 277 were clinically diagnosed (**Table 1**).

**Table 1.** Characteristics of study participants.

|  | **PSP (n = 1,718)** |  | **Control (n = 2,944)** |
| --- | --- | --- | --- |
|  | **Autopsy Confirmed**<br>**(n = 1,441)** | **Clinical Diagnosed**<br>**(n = 277)** |  |
| **Female** | 625 (43%) | 129 (46%) | 1,775 (60%) |
| **Age, y (SD)** | 68.38 (8.22) | 65.72 (7.68) | 81.19 (6.01) |
| **APOE $\epsilon 4^a$** |  |  |  |
| *$\epsilon 4$ carriers* | 350 (24%) | 57 (21%) | 905 (32%) |
| *Non-$\epsilon 4$ carriers* | 1,085 (75%) | 216 (78%) | 1,913 (65%) |
| *Data missing* | 6 (0.42%) | 4 (1%) | 126 (4%) |
| **APOE $\epsilon 2^b$** |  |  |  |
| *$\epsilon 2$ carriers* | 234 (16%) | 36 (13%) | 220 (8%) |
| *Non-$\epsilon 2$ carriers* | 1,193 (83%) | 238 (86%) | 2,522 (86%) |
| *Data missing* | 14 (1%) | 3 (1%) | 202 (7%) |
| **H2<sup>c</sup>** |  |  |  |
| *H2 carriers* | 158 (11%) | 27 (10%) | 1,182 (40%) |
| *Non-H2 carriers* | 1,283 (89%) | 250 (90%) | 1,761 (60) |
| *Data missing* | 0 (0%) | 0 (0%) | 1 (0.03%) |

<sup>a</sup>APOE $\epsilon 4$ is represented by the genotypes of rs429358-C.
<sup>b</sup>APOE $\epsilon 2$ is represented by the genotypes of rs7412-T.
<sup>c</sup>H2 haplotype is determined by the genotypes of rs8070723-G.
SD, standard deviation.

Considering that our sample set incorporated external controls from ADSP, initially collected for Alzheimer’s Disease (AD) studies, there was a potential selection biases for *APOE* ε4 and ε2 in controls. To rigorously validate our findings linked to *APOE*, we broke down the allele frequencies of *APOE* ε4 and ε2 by cohorts (**Table S2**), reviewed the study design of each cohort, and created an additional sample set by excluding those cohorts with selection bias against *APOE* ε4 or ε2 (**Supplementary Methods**).

### SNVs/indels quality controls

Only biallelic variants were included in common (Minor Allele Frequency [MAF] > 0.01) SNVs/indels analysis. Variants were removed if they were monomorphic, did not pass variant quality score recalibration (VQSR), had an average read depth ≥ 500, or if all calls have alignment depth (DP < 10) and genotype quality (GQ < 20). Individual calls with DP < 10 or GQ < 20 were set to missing. Indels were left aligned using the GRCh38 reference [20,21]. Common variants with a missing rate < 0.1, 0.25 < allele balance for heterozygous calls (ABHet) < 0.75, and Hardy-Weiberg Equilibrium tests (HWE) in controls > 1 × 10^-5^ were kept for analysis, leaving 7,945,112 SNVs/indels for analysis. Similar quality control procedures were applied to rare variants (**Supplementary Methods**). Then, we calculated the heritability of PSP using GCTA-LDMS [22] for common SNVs/indels (MAF > 0.01) and common plus rare SNVs/indels. A prevalence of 5 PSP cases per 100,000 individuals (0.00005) was used in the GCTA-LDMS analysis.

### Common SNVs/indels analysis

For association analysis, linear mixed model implemented in R Genesis [23] were used. Genetic relatedness matrix was obtained using KING [24]. PCs were obtained by PC-AiR [25] which accounts for sample relatedness. Sex and PC1-5 were adjusted in the linear mixed model. Age was not adjusted as more than half (1,159 of 1,718) of PSP cases had age missing. SNVs and indels with a *P* < 1 × 10^-6^ were reported along with the WGS quality metrics, such as QualByDepth (QD) and FisherStrand (FS), (**Table S3**).

For H1/H2 region, fine-mapping were analyzed using SuSie [26]. We ran the analysis several times assuming the number of maximum causal variants were from 2 to 10. The only variant (rs242561) robust to the choice of maximum causal variants was reported. To avoid potential confounding effects (particularly for *APOE* alleles), we also performed association analysis (**Table S4**, **Table S5**) for suggestive and genome-wide significant signals when excluding subjects from the three cohorts with selection bias against *APOE* alleles (ADSP-FUS1-APOEextremes, ADSP-FUS1-StEPAD1, and CacheCounty) along with cohorts with less than 10 subjects (NACC-Genentech, FASe-Families-WGS, and KnightADRC-WGS) (**Table S2**). We also performed additional experimental validation using TaqMan assay/Sanger sequencing to confirm the genotype of *APOE* observed from WGS (**Supplementary Methods**, **Table S6**).

### Rare SNVs/indels analysis

For aggregated tests of rare variants, we considered rare protein truncating variants (PTVs) and PTVs/damaging missense variants. Variant were annotated with ANNOVAR (version 2020-06-07) [27] and Variant Effect Predictor (VEP, version 104.3) [28]. PTVs were in protein coding genes (Ensembl version 104) [29] and had VEP consequence as stop gained, splice acceptor, splice donor or frameshift. Damaging missense variants were in protein coding genes (Ensembl version 104) and had a VEP consequence as missense, CADD score ≥ 15, and PolyPhen-2 HDIV of probably damaging. Rare variants were selected based on a MAF < 0.01% from gnomAD and a MAF < 1% in our dataset. The number of alternative allele variants in PTVs and PTVs/damaging missense variants was similar across sequencing centers and when evaluated for loss of function intolerant genes (observed/expected score upper confidence interval < 0.35 [30]) (**Fig. S14**) We performed SKAT-O and gene burden testing (SKATBinary, method=’burden’) for PTVs and PTV/damaging missense variants (**Supplementary Methods**). We also considered only PTVs or PTVs/missense variants in loss of function intolerant genes (observed/expected score upper confidence interval < 0.35[30]) when performing the tests. *P*-values were FDR corrected for the number of genes with a total minor allele count (MAC) ≥ 10. As SKAT-O does not calculate an odds ratio, we calculated the odds ratio of significant genes using logistic regression with the same covariates as SKAT-O and burden testing, and the same variant weights.

We evaluated the C1 module, a gene set, which was previously shown to be composed of neuronal genes and enriched for common variants in PSP [31]. We performed a permutation test (N=1000) of random gene set modules from brain expressed genes that contained the same number of genes as C1. From the human protein atlas (www.proteinatlas.org) [32], brain expressed genes were defined as the union of unique proteins from the cerebral cortex, basal ganglia and midbrain (N=15,638). We calculated SKAT-O *P*-values from these random gene modules to determine the null distribution. We calculated the unadjusted odds ratio of significant genes or gene sets by summing the number of alternate alleles in the gene set among the total number alleles in cases and controls. Normalized quantification (TPM) gene expression across tissues was obtained from Genotype-Tissue Expression (GTEx) [33]. The expression of ZNF592 and C1 module (summarized as an eigengene [34]) were plotted.

### SV detection and filtering

For each sample, SVs were called by Manta (v1.6.0) [35] and Smoove (v0.2.5) [36] with default parameters. Calls from Manta and Smoove were merged by Svimmer [37] to generate a union of two call sets for a sample. Then, all individual sample VCF files were merged together by Svimmer as input to Graphtyper2 (v2.7.3) [37] for joint genotyping. SV calls after joint-genotyping are comparable across the samples, therefore, can be used directly in genome-wide association analysis [37]. A subset of SV calls was defined as high-quality calls [37]. Details of SV calling pipeline were in our previous study [38].For each individual SV reported, Samplot [39] or IGV [40] were used to keep only high-confident CNVs and inversions that are supported by read depth or split reads; for insertions, we kept high-confident insertions that are high-quality and not in the masked regions (**Supplementary Methods**).

### SV analysis

For SV association, more strict sample filtering was applied: outlier samples with too many (larger than median + 4*MAD) CNV/insertion calls or too little (smaller than median - 4*MAD) high-quality CNV/insertion calls were removed. There were 4,432 samples (1,703 cases and 2,729 controls) remaining for PSP SV association analysis. Due to more false positives being picked up, the genomic inflation would be high (λ = 1.89, **Fig. S9**) if all SVs were included in the analysis. Therefore, we restricted our analysis to high-quality SVs only, making the genomic inflation drop to 1.27 (**Fig. S9**). The 14,792 high-quality common SVs (MAF > 0.1) with call rate > 0.5 were included in the analysis. Mixed model implemented in R Genesis were used for association. Sex, PCR information, SV PCs 1-5, and SNV PCs 1-5 were adjusted in the mixed model. After association, we manually inspect deletions, duplications, and inversions by Samplot or IGV to keep only those with support from read depth, split read or insert size. For insertions, those not on masked regions were reported.

For SVs inside the H1/H2 region, all SVs those that are not high-quality are included. Then, we removed SVs with missing rate > 0.5 and manual inspect deletions, duplications, and inversions by Samplot or IGV to keep only those with support from read depth, split read or insert size. For insertions, those high-quality ones not on masked regions were kept for analysis. LD between SVs was calculated using PLINK (V1.90 beta) [41].Rare SV burden on H1/H2 region was evaluated by SKAT-O [42] adjusting for gender and PCs 1-5. As SKAT-O does not calculate an odds ratio, we calculated the odds ratio using logistic regression with the same covariates.

## Results

### Common SNVs and indels associated with PSP

We conducted whole genome sequencing at 30x coverage in 4,662 European-ancestry samples (1,718 individuals with PSP of which 1,441 were autopsy confirmed and 277 were clinically diagnosed and 2,944 control subjects, **Table 1**). We successfully replicated the association of known loci at *MAPT*, *MOBP* and *STX6* [8,12,13] and identified a novel signal in *APOE* with a genome-wide significance of *P* < 5 × 10^-8^ (**Fig. 1, Fig. S1, Table 2, Table S3**). Furthermore, eight loci showed suggestive significance (5 × 10 ^-8^ < *P* < 1 × 10^-6^), including two loci reported genome-wide significant (*SLCO1A2* and *DUSP10*) and one locus (*SP1*) reported suggestive significant in previous studies [12,13], as well as five new loci in *FCHO1*/*MAP1S*, *KIF13A*, *TRIM24*, *ELOVL1* and *TNXB*.

**Fig. 1:**
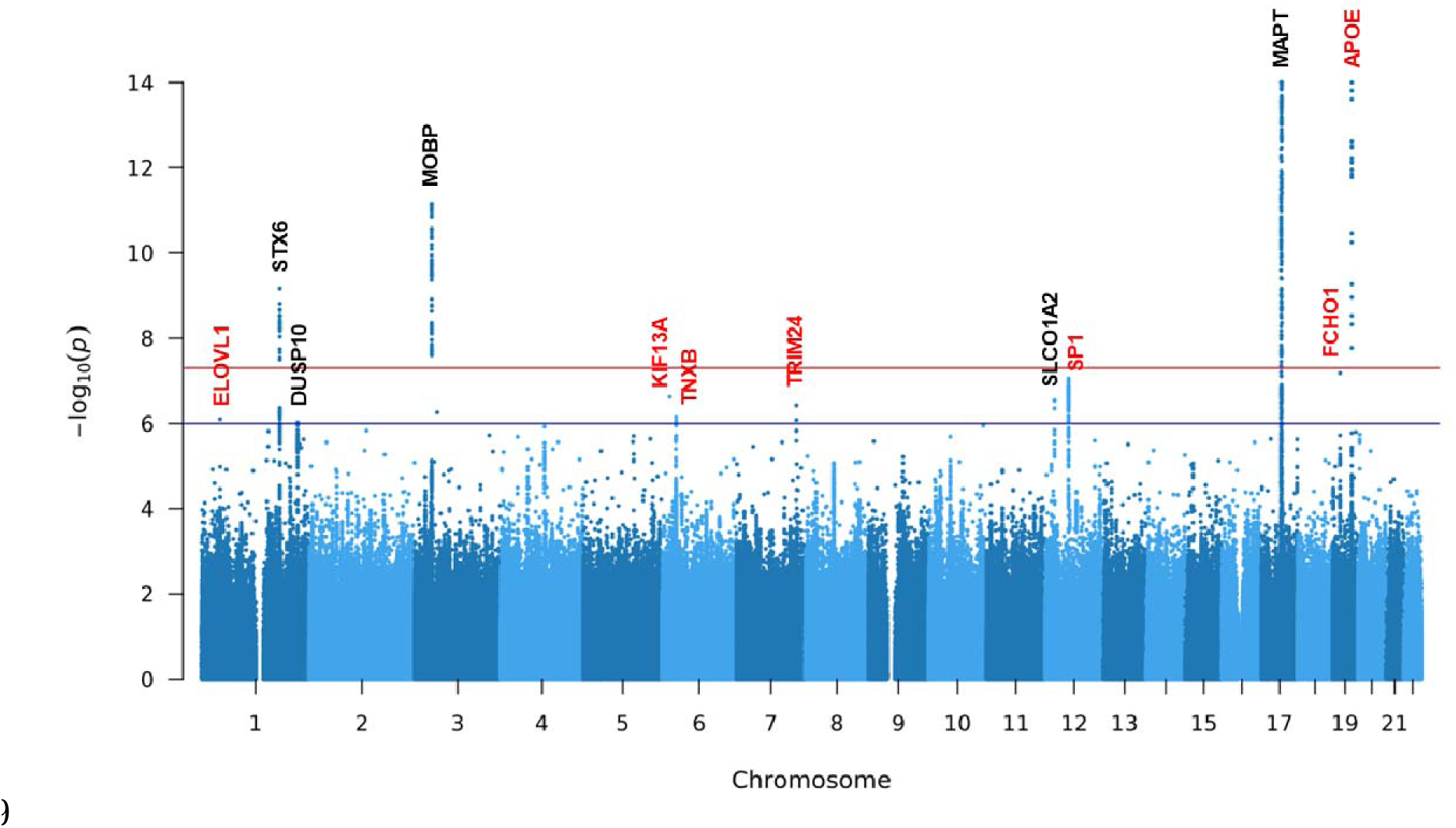
Manhattan plot of SNVs/indels for PSP. Loci with a suggestive or genome-wide significant signal are annotated (novel loci in red and known loci in black). Variants with a *P*-value below 1 × 10^-14^ are not shown. The red horizontal line represents genome-wide significance level (5 × 10^-8^). The blue horizontal line represents suggestive significance level (1 × 10^-6^).

**Table 2.**
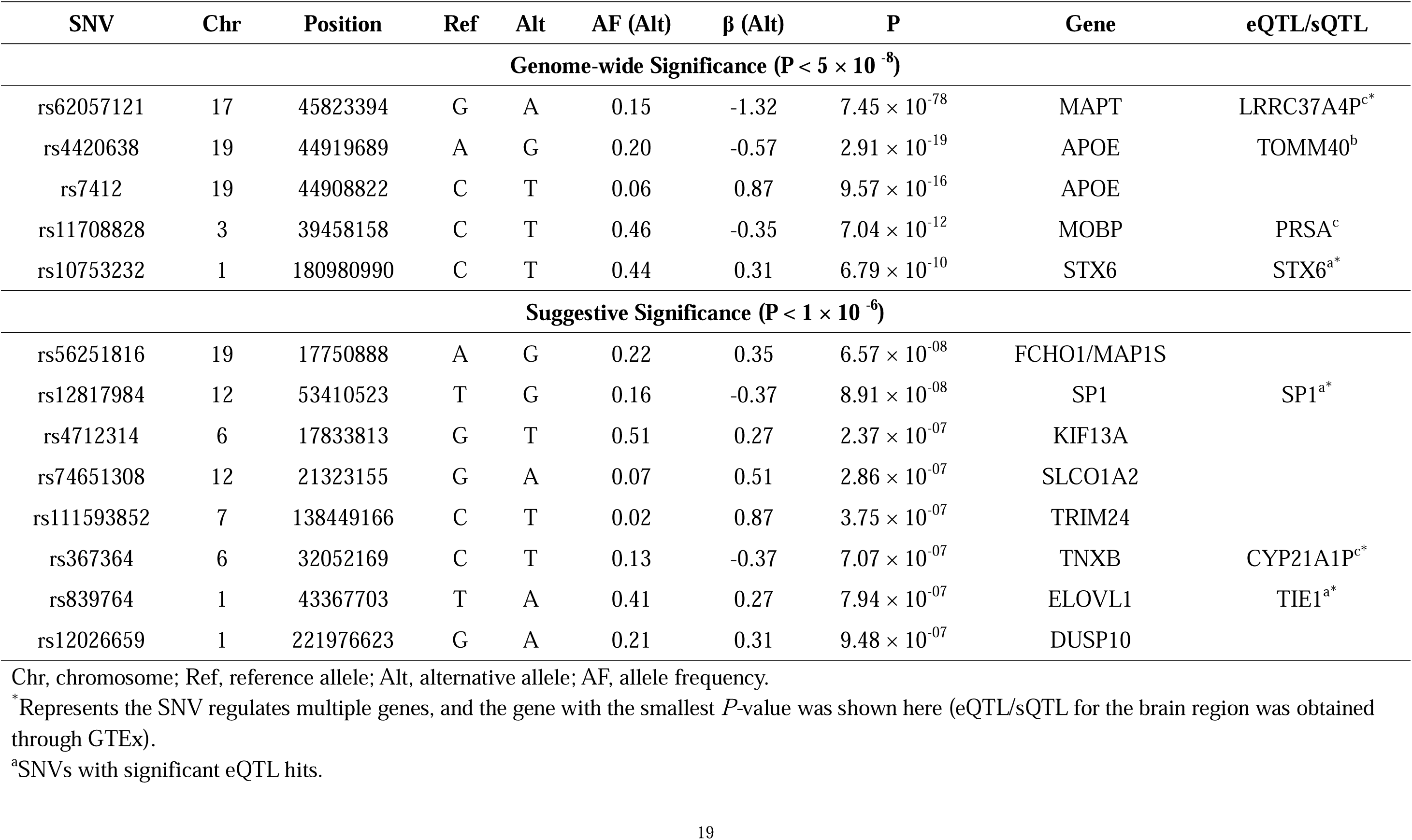
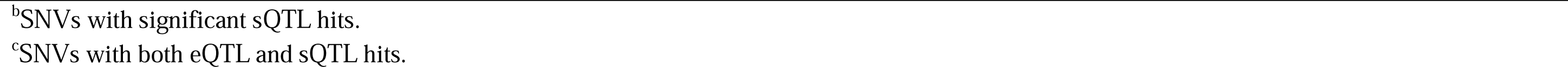
Genome-wide and suggestive significant loci.

| SNV | Chr | Position | Ref | Alt | AF (Alt) | $\beta$ (Alt) | P | Gene | eQTL/sQTL |
| --- | --- | --- | --- | --- | --- | --- | --- | --- | --- |
| **Genome-wide Significance ($P < 5 \times 10^{-8}$)** |  |  |  |  |  |  |  |  |  |
| rs62057121 | 17 | 45823394 | G | A | 0.15 | -1.32 | $7.45 \times 10^{-78}$ | MAPT | LRRC37A4P<sup>c*</sup> |
| rs4420638 | 19 | 44919689 | A | G | 0.20 | -0.57 | $2.91 \times 10^{-19}$ | APOE | TOMM40<sup>b</sup> |
| rs7412 | 19 | 44908822 | C | T | 0.06 | 0.87 | $9.57 \times 10^{-16}$ | APOE | |
| rs11708828 | 3 | 39458158 | C | T | 0.46 | -0.35 | $7.04 \times 10^{-12}$ | MOBP | PRSA<sup>c</sup> |
| rs10753232 | 1 | 180980990 | C | T | 0.44 | 0.31 | $6.79 \times 10^{-10}$ | STX6 | STX6<sup>a*</sup> |
| **Suggestive Significance ($P < 1 \times 10^{-6}$)** |  |  |  |  |  |  |  |  |  |
| rs56251816 | 19 | 17750888 | A | G | 0.22 | 0.35 | $6.57 \times 10^{-08}$ | FCHO1/MAP1S | |
| rs12817984 | 12 | 53410523 | T | G | 0.16 | -0.37 | $8.91 \times 10^{-08}$ | SP1 | SP1<sup>a*</sup> |
| rs4712314 | 6 | 17833813 | G | T | 0.51 | 0.27 | $2.37 \times 10^{-07}$ | KIF13A | |
| rs74651308 | 12 | 21323155 | G | A | 0.07 | 0.51 | $2.86 \times 10^{-07}$ | SLCO1A2 | |
| rs111593852 | 7 | 138449166 | C | T | 0.02 | 0.87 | $3.75 \times 10^{-07}$ | TRIM24 | |
| rs367364 | 6 | 32052169 | C | T | 0.13 | -0.37 | $7.07 \times 10^{-07}$ | TNXB | CYP21A1P<sup>c*</sup> |
| rs839764 | 1 | 43367703 | T | A | 0.41 | 0.27 | $7.94 \times 10^{-07}$ | ELOVL1 | TIE1<sup>a*</sup> |
| rs12026659 | 1 | 221976623 | G | A | 0.21 | 0.31 | $9.48 \times 10^{-07}$ | DUSP10 | |

Chr, chromosome; Ref, reference allele; Alt, alternative allele; AF, allele frequency.
\*Represents the SNV regulates multiple genes, and the gene with the smallest *P*-value was shown here (eQTL/sQTL for the brain region was obtained through GTEx).
<sup>a</sup>SNVs with significant eQTL hits.
<sup>b</sup>SNVs with significant sQTL hits.
<sup>c</sup>SNVs with both eQTL and sQTL hits.

### MAPT, MOBP and STX6

In the *MAPT* region, a multitude of SNVs and indels in high linkage disequilibrium (LD) with the H1/H2 haplotype remains the most significant association with PSP (**Fig. S2A**). From our analysis, the prominent signal within the *MAPT* region is rs62057121 (*P* = 7.45 × 10^-78^, β = −1.32, MAF = 0.15). Fine mapping suggests that rs242561 (*P* = 4.49 × 10^-74^, β = −1.23, MAF = 0.16) is likely to be a causal SNV underling the statistical significance. The SNP rs242561 is located in an enhancer region, containing an antioxidant response element that binds with NRF2/sMAF protein complex. The T allele of rs242561 showed a stronger binding affinity for NRF2/sMAF in ChIP-seq analysis, therefore inducing a significantly higher transactivation of the *MAPT* gene [43]. rs242561 and rs62057151 were both in high LD (r^2^ > 0.9) with H1/H2 (defined by the 238 bp deletion in *MAPT* intron 9) and represented the same association signal as the H1/H2. However, in previous studies [8,44], the H1c tagging SNV (rs242557) inside the H1/H2 region was found to be significant when conditioning on H1/H2. We confirmed that rs242557 was genome-wide significant after adjusting for H1/H2 (*P* = 3.68 × 10^-15^, β = 0.39, MAF = 0.42) though in weak LD with H1/H2 (r^2^ = 0.14). To pinpoint the causal genes underlying the association in H1/H2 requires additional functional study. For example, Cooper *et al.* [11] analyzed transcriptional regulatory activity of SNVs and suggested *PLEKHM1* and *KANSL1* were probable causal genes in H1/H2 besides *MAPT*. In *MOBP* (rs11708828, *P* = 7.04 × 10^-12^, β = −0.35, MAF = 0.46, **Fig. S2B**) and *STX6* (rs10753232, *P* = 6.79 × 10^-10^, β = 0.31, MAF = 0.44, **Fig. S2C**), the associated variants were of high allele frequency and exhibited moderate effect size.

### APOE and risk of PSP

One newly identified significant locus from our analysis is the well-known AD risk gene, *APOE*. We observed a significant association between the *APOE* ε2 haplotype and an elevated risk of PSP (*P* = 9.57 × 10^-16^, β = 0.87, MAF = 0.06, **Table 3**, **Fig. S3B**). The *APOE* ε2 haplotype is encoded by rs429358-T and rs4712-T, which is considered a protective allele in AD. The increased risk of *APOE* ε2 in PSP has been previously reported in a Japanese cohort, albeit with a relatively small sample size [45]. Furthermore, Zhao *et al*. [46] confirmed that *APOE* ε2 is linked to increased tau pathology in the brains of individuals with PSP and reported a higher frequency of homozygosity of *APOE* ε2 in PSP with an odds ratio of 4.41. Consistent with these findings, our dataset exhibited a higher frequency of homozygosity of rs7412-T in PSP, yielding an odds ratio of 3.91.

**Table 3.** Allele Frequency of *APOE* _ε_**4 SNV (rs429358) and** _ε_**2 SNV (rs7412)**

| Studies | rs429358 |  | rs7412 |  |
| --- | --- | --- | --- | --- |
|  | AF (Case) | AF (Control) | AF (Case) | AF (Control) |
| PSP WGS (This study) | 0.1279 | 0.1742 | 0.0844 | 0.0414 |
| PSP GWAS [52] | 0.1159 | 0.1366 | 0.0826 | 0.0794 |
| 1000 Genomes Project [18] |  | 0.1512 |  | 0.0771 |
| ExAC<br>European (non-Finnish) [53] |  | 0.2078 |  | 0.1060 |
| gnomAD V4<br>European (non-Finnish) [54] |  | 0.1506 |  | 0.0783 |
| TOPMed Freeze 8<br>NFE (Non-Finnish European) |  | 0.1501 |  | 0.0752 |
| ADSP R3 Non-Hispanic<br>White [55] | 0.3139<br>(AD as cases) | 0.1803 | 0.0244<br>(AD as cases) | 0.0406 |

For *APOE* ε4 allele, contrary to its association with AD, we observed that rs429358-C exhibits a protective effect against PSP (*P* = 5.71 × 10^-18^, β = −0.60, MAF = 0.16, **Table 3**). The lead SNV demonstrating this protective association from our analysis is rs4420638 (*P* = 2.91 × 10^-19^, β = −0.57, MAF = 0.20, **Fig. S3A**), which is in LD (r^2^ = 0.74) with rs429358. In a previous PSP GWAS conducted by Hoglinger *et al*. [8], another *APOE* ε4 tagging SNV (rs2075650, r^2^ = 0.52 with rs429358) was also found to be diminished (MAF_case = 0.11 and MAF_control = 0.15) in PSP, although not reaching significance (*P* = 1.28 × 10^-5^). Notably, in our analysis, rs2075650 reached genome-wide significance (*P* = 3.39 × 10^-13^, β = −0.51, MAF = 0.15). *APOE* ε4 or ε2 displayed an independent effect for PSP risk without a significant epistatic interaction with H1/H2 haplotype (*P* > 0.05) (**Fig. S4**).

Given that our dataset included external controls from ADSP collected for Alzheimer’s disease studies, there were a potential selection biases for *APOE* ε4 and ε2 in controls. To address this concern, we broke down the allele frequencies of *APOE* ε4 and ε2 by cohorts (**Table S2**) and indicated cohorts with potential selection bias. The association analysis excluding these cohorts shows the ε2 SNV (rs7412, *P* = 1.23 × 10^-12^, β = 0.70, MAF = 0.06) remained genome-wide significant and ε4 SNV (rs429358, *P* = 0.02, β = −0.16, MAF = 0.14) was nominal significant (**Table S4**, **Table S5**).

### Suggestive significant loci

Eight loci were suggestive of significance in our analysis of which three, *SLCO1A2*, *DUSP10*, and *SP1*, were previously reported [12,13]. In *SLCO1A2*, the lead SNV rs74651308 (*P* = 2.86 × 10^-7^, β = 0.51, MAF = 0.07, **Fig. S5A**) is intronic and in LD (r^2^ = 0.98) with missense SNV rs11568563 (*P* = 1.45 × 10^-6^, β = 0.47, MAF = 0.07), which was reported in a previous study [12]. About 250 kb upstream of *DUSP10* lies the previously reported SNV rs6687758 [12] (*P* = 3.36 × 10^-6^, β = 0.29, MAF = 0.21), which is in LD (r^2^ = 0.98) with the lead SNV rs12026659 in our analysis (*P* = 9.48 × 10^-7^, β = 0.31, MAF = 0.21, **Fig. S5B**). In *SP1*, the reported indel rs147124286 [13] (*P* = 4.39 × 10^-7^, β = −0.35, MAF = 0.16) is in LD (r^2^ = 0.995) with the lead SNV rs12817984 (*P* = 8.91 × 10^-8^, β = −0.37, MAF = 0.16, **Fig. S5C**). Notably, disruption of a transcriptional network centered on *SP1* by causal variants has been implicated previously in PSP [11].

Five newly discovered suggestive loci are in *FCHO1/MAP1S, KIF13A, TRIM24, TNXB,* and *ELOVL1*. Within *FCHO1*/*MAP1S*, the most significant signal (rs56251816, *P* = 6.57 × 10^-8^, β = 0.35, MAF = 0.22, **Fig. S6A**) is in the intron of *FCHO1*. rs56251816 is a significant expression quantitative trait locus (eQTL) for both *FCHO1* and *MAP1S* (13 kb upstream of *FCHO1*) in the Genotype-Tissue Expression (GTEx) project [47]. *MAP1S* encodes a microtubule associated protein that is involved in microtubule bundle formation, aggregation of mitochondria and autophagy [48], and therefore, is more relevant than *FCHO1* regarding PSP. *KIF13A*, which encodes a microtubule-based motor protein was also suggestive of significance (rs4712314, *P* = 2.37 × 10^-7^, β = 0.27, AF = 0.51, **Fig. S6B**). The significance in genes involved in microtubule-based processes, such as *MAPT*, *MAP1S* and *KIF13A*, implicates the neuronal cytoskeleton as a convergent aspect of PSP etiology.

Other variants with suggestive association evidence include *TRIM24* (rs111593852, *P* = 3.75 × 10^-7^, β = 0.87, MAF = 0.02, **Fig. S7A**). *TRIM24* is involved in transcriptional initiation and shows differential expression in individuals with Parkinson disease [49,50]. Another suggestive locus is *TNXB*, located in the major histocompatibility complex (MHC) region on chromosome 6, with the lead SNV rs367364 (*P* = 7.07 × 10^-7^, β = −0.37, MAF = 0.13, **Fig. S7B**). Finally, *ELOVL1* yields suggestive evidence of association (rs839764, *P* = 7.94 × 10^-7^, β = 0.27, MAF = 0.41, **Fig. S7C**).

This gene encodes an enzyme that elongates fatty acids and can cause a neurological disorder with ichthyotic keratoderma, spasticity, hypomyelination and dysmorphic features [51]. Furthermore, we found a few SNV/indels that reached genome-wide or suggestive significance without other supporting variants in LD (**Fig.S1**, **Table S3**). These signals could be due to sequencing errors and need further experimental validation.

### Rare SNVs/indels and network analysis

The heritability of PSP for common SNVs and indels (MAF > 0.01) was estimated to be 20%, while common plus rare SNVs/indels was estimated to be 23% from our analysis using GCTA-LDMS [22]. Therefore, we performed aggregated tests for rare SNVs and indels, and identified *ZNF592* (SKAT-O FDR=0.043, burden test FDR=0.041) with an of OR = 1.08 (95% CI: 1.008-1.16) (**Fig. 2**, **Table 4**, **Table S7**) for protein truncating or damaging missense variants. There was no genomic inflation with a λ=1.07 (**Fig. 2**). Risk in *ZNF592* was imparted by 16 unique variants, with one splice donor and 15 damaging missense variants (**Table S7**). *ZNF592* has not been previously associated with PSP but showed moderate RNA expression in the cerebellum compared to other tissues from GTEx (**Fig. S8**). There were no significant genes identified when evaluating PTVs only or when restricting to loss of function intolerant genes.

**Fig. 2:**
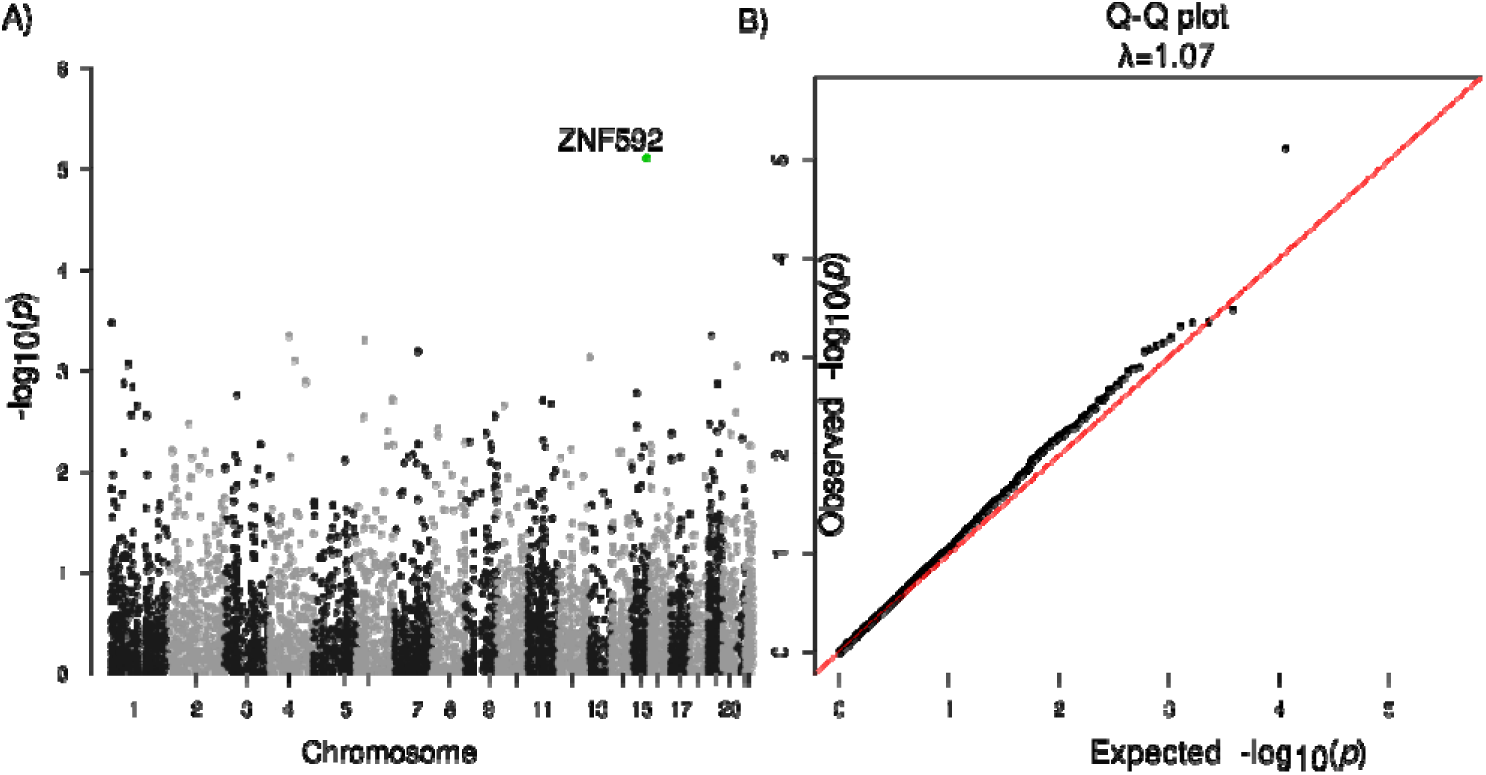
Association analysis of rare SNVs/indels. **A.** Manhattan plot for genes with protein truncating variants or damaging missense variants. **B.** Q-Q plot of gene *P*-values with protein truncating variants or damaging missense variants.

**Table 4.**
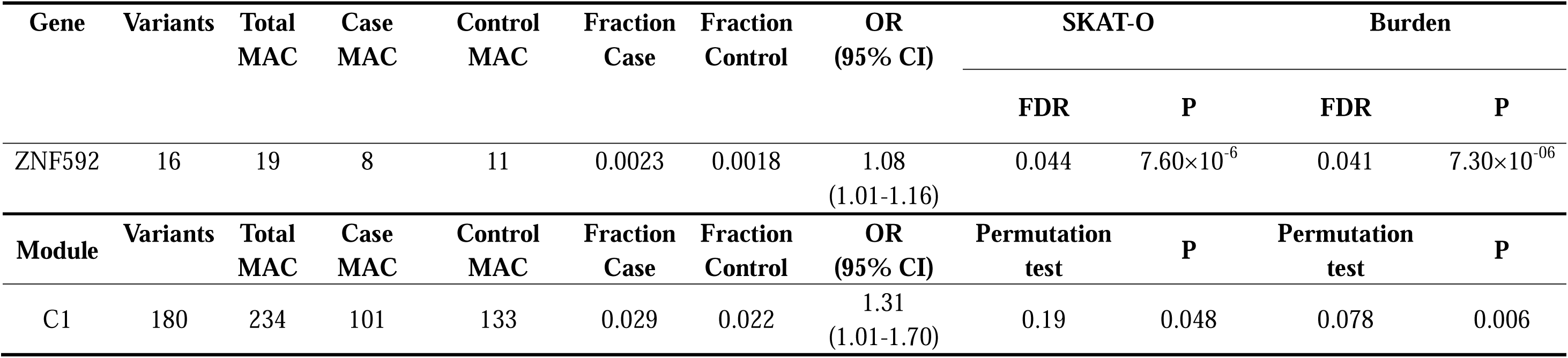
Association analysis of ZNF592 and the C1 module.

Considering that genes do not operate along, but rather within signaling pathways and networks, we and others have shown that better understanding of disease mechanisms can be achieved through gene network analysis [56–58]. Therefore, we scrutinized rare variants within a network framework, focusing on co-expression network analysis performed in PSP post mortem brain that had previously identified a brain co-expression module, C1, which was conserved at the protein interaction level and enriched for common variants in PSP [31]. We found this C1 neuronal module was significantly enriched with PSP rare variants (*P* = 0.006, OR [95% CI] = 1.31 [1.01-1.70], **Table 4**; **Table S8**). Genes from the C1 module were more likely to be loss of function intolerant compared to the background of all brain expressed genes (**Fig. S8**). To ensure that this was association not spurious, we performed permutation testing using random gene modules of brain expressed genes with the same number of genes as C1. The C1 module remains significant (Permutation *P* = 0.078). Exploring GTEx, we found that C1 genes are highly expressed in brain tissues including the cerebellum, frontal cortex, and basal ganglia (**Fig. S8**), consistent with regions affected in this disorder.

### SVs associated with PSP

Seven high-confident SVs achieved genome-wide significance with PSP (**Table 5**, **Fig. S9**), including three deletions tagging the H2 haplotype. The most significant signal is a 238 bp deletion in *MAPT* intron 9 (**Fig. S10A**, chr17:46009357-46009595, *P* = 3.14 × 10^-50^, AF = 0.16) that has been reported on the H2 haplotype [59,60] and is in LD (r^2^ = 0.99) with the lead SNV, rs62057121 (chr17:45823394, *P* = 7.45 × 10^-78^, β = −1.32, MAF = 0.15), in the *MAPT* region. Adding to this, two other deletions, one spanning 314 bp (**Fig. S10B**, chr17:46146541-46146855, AF = 0.19) and the other covering 323 bp (**Fig. S10C**, chr17:46099028-46099351, AF = 0.22), both are Alu elements and in LD (r^2^ > 0.8) with the top signal (the 238 bp deletion). This observation indicates that transposable elements may play an important role in the evolution of H1/H2 haplotype structure.

**Table 5.**
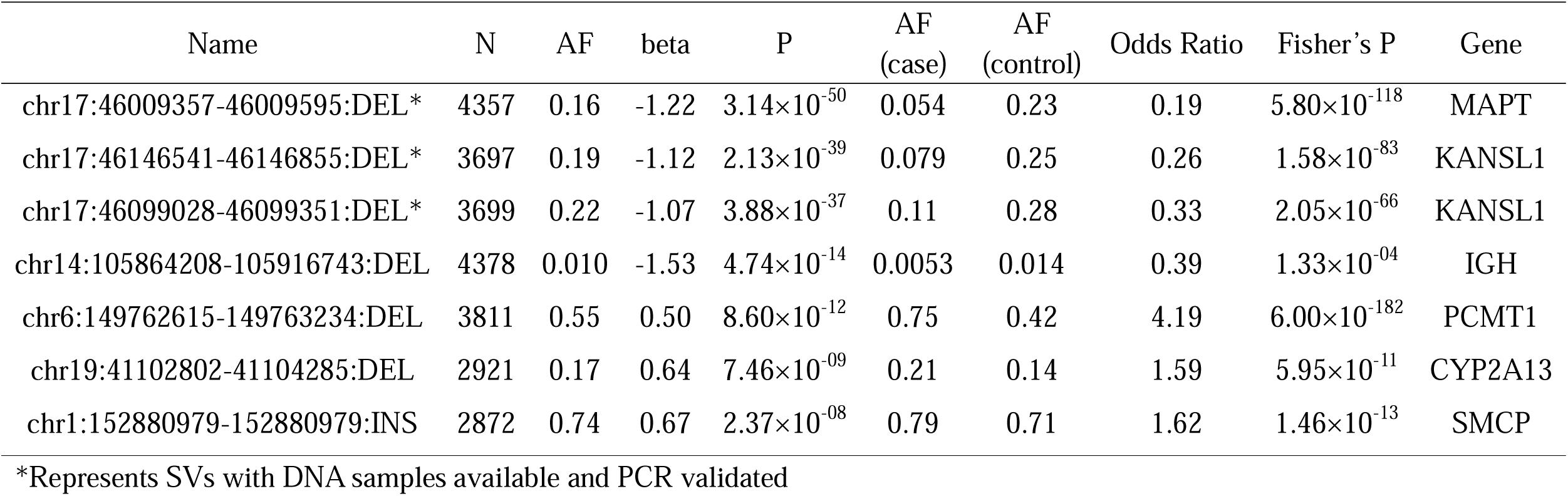
Significant structural variants from association analysis (P < 5 × 10^-8^).

Beyond the identified SVs in the H1/H2 region, we uncovered a significant deletion (chr14:105864208-105916743, *P* = 4.74 × 10^-14^, AF = 0.01) within the immunoglobulin heavy locus (*IGH*), which is a complex SV region (**Fig. S11**) related to antigen recognition. Moreover, a 619 bp deletion (chr6:149762615-149763234, *P* = 8.60 × 10^-12^, AF = 0.55; **Fig. S10D**) in *PCMT1* displayed increased risk of PSP with an odds ratio of 4.19. The odds ratio increased to 8.38 when comparing 1,244 individuals with homozygous deletions in *PCMT1* with the rest of sample set. *PCMT1* encodes a type □ class of protein carboxyl methyltransferase enzyme that is highly expressed in the brain [61] and is able to ameliorate Aβ_25-35_ induced neuronal apoptosis [62,63]. Additionally, we found a deletion between *CYP2F1* and *CYP2A13* (chr19:41102802-41104285, AF = 0.17) and an insertion in *SMCP* (chr1:152880979-152880979, AF = 0.74) which were also significant (**Table 5**). The 1.5 kb deletion (chr19:41102802-41104285) almost completely overlaps the SINE-VNTR-Alus (SVA) transposon region annotated by RepeatMasker [64].

### SVs in H1/H2 haplotype region

The H1/H2 region stands out as the pivotal genetic risk factor for PSP [8,65]. The H2 haplotype exhibits a reduced odds ratio of 0.19, as we observed the allele frequency of the 238 bp H2-tagging deletion is 23% in PSP and only 5% in control (*P* < 2.2 × 10^-16^). Moreover, our analysis pointed out five common (MAF > 0.01) and 12 rare deletions and duplications in the region (**Table 6**), ranging from 88 bp to 47 kb. Additionally, one common and four rare high-confidence insertions were reported in the region.

**Table 6.**
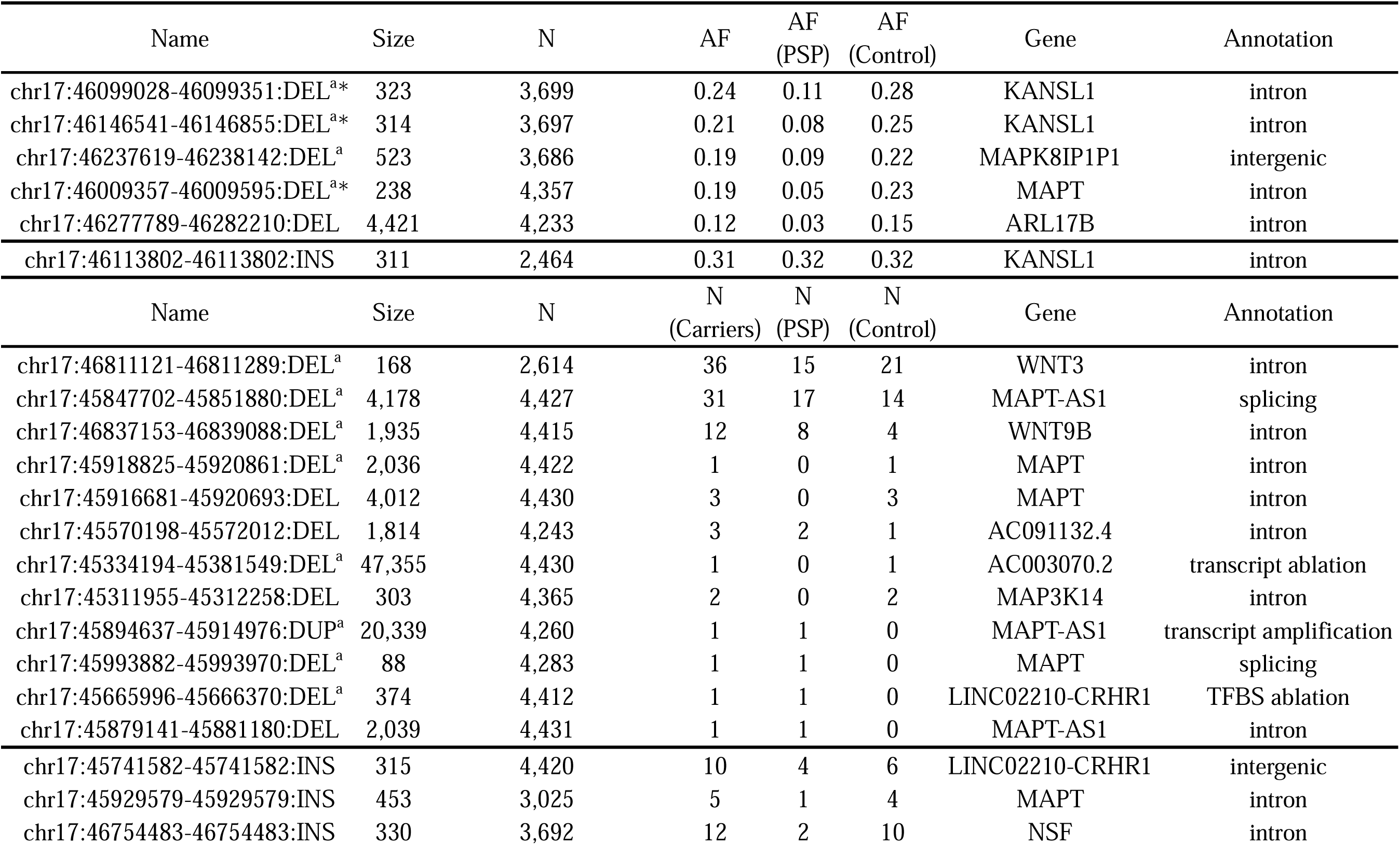
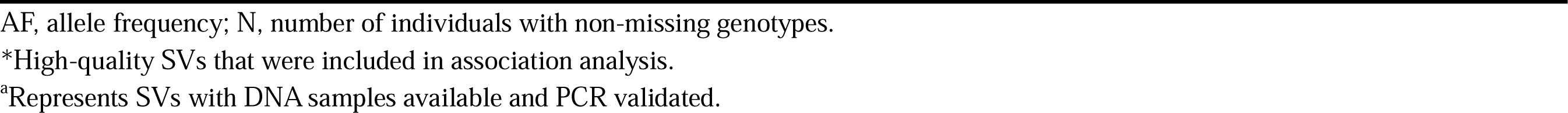
High-confident structural variants in the H1/H2 haplotype region.

| Name | Size | N | AF | AF<br>(PSP) | AF<br>(Control) | Gene | Annotation |
| --- | --- | --- | --- | --- | --- | --- | --- |
| chr17:46099028-46099351:DEL<sup>a*</sup> | 323 | 3,699 | 0.24 | 0.11 | 0.28 | KANSL1 | intron |
| chr17:46146541-46146855:DEL<sup>a*</sup> | 314 | 3,697 | 0.21 | 0.08 | 0.25 | KANSL1 | intron |
| chr17:46237619-46238142:DEL<sup>a</sup> | 523 | 3,686 | 0.19 | 0.09 | 0.22 | MAPK8IP1P1 | intergenic |
| chr17:46009357-46009595:DEL<sup>a*</sup> | 238 | 4,357 | 0.19 | 0.05 | 0.23 | MAPT | intron |
| chr17:46277789-46282210:DEL | 4,421 | 4,233 | 0.12 | 0.03 | 0.15 | ARL17B | intron |
| chr17:46113802-46113802:INS | 311 | 2,464 | 0.31 | 0.32 | 0.32 | KANSL1 | intron |
| Name | Size | N | N<br>(Carriers) | N<br>(PSP) | N<br>(Control) | Gene | Annotation |
| chr17:46811121-46811289:DEL<sup>a</sup> | 168 | 2,614 | 36 | 15 | 21 | WNT3 | intron |
| chr17:45847702-45851880:DEL<sup>a</sup> | 4,178 | 4,427 | 31 | 17 | 14 | MAPT-AS1 | splicing |
| chr17:46837153-46839088:DEL<sup>a</sup> | 1,935 | 4,415 | 12 | 8 | 4 | WNT9B | intron |
| chr17:45918825-45920861:DEL<sup>a</sup> | 2,036 | 4,422 | 1 | 0 | 1 | MAPT | intron |
| chr17:45916681-45920693:DEL | 4,012 | 4,430 | 3 | 0 | 3 | MAPT | intron |
| chr17:45570198-45572012:DEL | 1,814 | 4,243 | 3 | 2 | 1 | AC091132.4 | intron |
| chr17:45334194-45381549:DEL<sup>a</sup> | 47,355 | 4,430 | 1 | 0 | 1 | AC003070.2 | transcript ablation |
| chr17:45311955-45312258:DEL | 303 | 4,365 | 2 | 0 | 2 | MAP3K14 | intron |
| chr17:45894637-45914976:DUP<sup>a</sup> | 20,339 | 4,260 | 1 | 1 | 0 | MAPT-AS1 | transcript amplification |
| chr17:45993882-45993970:DEL<sup>a</sup> | 88 | 4,283 | 1 | 1 | 0 | MAPT | splicing |
| chr17:45665996-45666370:DEL<sup>a</sup> | 374 | 4,412 | 1 | 1 | 0 | LINC02210-CRHR1 | TFBS ablation |
| chr17:45879141-45881180:DEL | 2,039 | 4,431 | 1 | 1 | 0 | MAPT-AS1 | intron |
| chr17:45741582-45741582:INS | 315 | 4,420 | 10 | 4 | 6 | LINC02210-CRHR1 | intergenic |
| chr17:45929579-45929579:INS | 453 | 3,025 | 5 | 1 | 4 | MAPT | intron |
| chr17:46754483-46754483:INS | 330 | 3,692 | 12 | 2 | 10 | NSF | intron |

AF, allele frequency; N, number of individuals with non-missing genotypes.
\*High-quality SVs that were included in association analysis.
<sup>a</sup>Represents SVs with DNA samples available and PCR validated.

Of the five common deletions and duplications (**Fig. S12**), three show genome-wide significant association with the disease (**Table 2**); four are located in regions with transposable elements (SVA, L1, or Alu) and in LD (r^2^ from 0.63 to 0.92) with the 238 bp H2-tagging deletion. This further highlights the important role of transposable elements in shaping the landscape of H1/H2 region.

Among the 12 rare deletions and duplications (**Fig. S13**), five are located in potentially functional regions, such as splice sites, exons, and transcription factor binding sites (**Table 6**). Particularly, one deletion (chr17:45993882-45993970) in exon 9 of *MAPT* was identified in a PSP patient, adding to previous reports of exonic deletions in the *MAPT* in frontotemporal dementia, such as deletion of exon 10 [66] and exons 6-9 [67] in *MAPT*. Using the SKAT-O test (N = 4,432), the 12 rare CNVs displayed a significantly higher burden in PSP than controls (*P* = 0.01, OR = 1.64).

## Discussion

Through comprehensive analysis of whole genome sequence, we identified SNVs, indels and SVs contributing to the risk of PSP. For common SNVs, previously reported regions, including *MAPT*, *MOBP*, STX6, *SLCO1A2*, *DUSP10*, and *SP1* [8,12,13] were replicated in our analysis and novel loci in *APOE*, *FCHO1*/*MAP1S*, *KIF13A*, *TRIM24*, *ELOVL1*, and *TNXB* were discovered. *EIF2AK3* which was significantly associated with PSP in a previous GWAS [8] did not reach significance in our study. In the current study, the SNV with the lowest *P* around *EIF2AK3* was rs13003510 (*P* = 8.30 × 10^-5^, β = 0.22, MAF = 0.3).

The *APOE* ε4 haplotype was of particular interest as it is a common risk factor for AD, explaining more than a 1/3 of population attributable risk [68,69]. Typically, individuals with one copy of the *APOE* ε4 allele (rs429358-C and rs4712-G) have approximately a threefold increased risk of developing AD, while those with two copies of the allele have an approximately a 12-fold increase in risk [70]. In striking contrast, the ε4 tagging allele rs429358 was protective in PSP and the ε2 tagging allele rs7412 was deleterious. This observation is particularly intriguing since both AD and PSP have intracellular aggregated tau as a prominent neuropathologic feature. Notably, both ε2 allele and ε4 allele have been associated with tau pathology burden in the brain of mice models [46,71], which raises the question of distinct tau species in 4R-PSP versus 3R-4R-AD. It is also notable that the ε2 allele is also associated with increased risk for age-related macular degeneration (AMD), and the ε4 allele was associated with decreased risk [72,73]. These results demonstrate that the same variant may have opposite effects in different degenerative diseases. This is especially important, given the advent of gene editing as a therapeutic modality, and programs focused on changing *APOE* ε4 to ε2. Although this therapy would likely decrease risk for AD, our results indicate that it would increase risk for PSP, in addition to AMD. From this standpoint, caution is warranted in germ-line genome editing until the broad spectrum of phenotypes associated with human genetic variation is understood.

Burden association tests are an highly valuable for addressing sample size limitations in analyzing rare variants [74]. Indeed, burden testing allowed us to identify *ZNF592*, a classical C2H2 zinc finger protein (ZNF) [75,76], as a candidate risk gene. ZNF proteins have been causative or strongly associated with large numbers of neurodevelopmental disease [77,78] and neurodegenerative disease including Parkinson’s disease [79] and Alzheimer’s disease [80,81]. *ZNF592* was initially thought to be responsible for autosomal recessive spinocerebellar ataxia 5 from a consanguineous family with neurodevelopmental delay including cerebellar ataxia and intellectual disability due to a homozygous G1046R substitution [82]. However, further analysis of this family identified *WDR73* to be the most likely causative gene, consistent with Galloway-Mowat syndrome, although *ZNF592* may have contributed to the phenotype [83].

We also extended classical gene-based burden analysis to consider rare risk burden in the context of a gene set defined by co-expression networks [31,84]. We leveraged combined previous proteomic and transcriptomic analysis of post-mortem brain from patients afflicted with PSP, and showed that rare variants enrich in the C1 neuronal module, which was the same module enriched with common variants [31]. This, along with our recent work identifying a neuronally-enriched transcription factor network centered around SP1 disrupted by PSP common genetic risk, suggests that although PSP neuropathologically is defined by tufted astrocytes and oligodendroglial coiled bodies [6,85,86], initial causal drivers of PSP appear to be primarily neuronal.

In analysis of SVs, we found deletions in *PCMT1* and *IGH* were significantly associated with PSP. The *IGH* deletions are in a complex region on chromosome 14 that encodes immunoglobins recognizing foreign antigens. The size of the *IGH* deletion varies across individuals (**Fig. S9**). In addition, the *IGH* deletions can be accompanied by other deletions, duplications, and inversions (**Fig. S9**). These combined make the experimental validation of the deletion challenging. The *PCMT1* deletion is common (AF = 0.55) with an odds ratio of 8.38 for PSP in homozygous individuals.

There were limitations to this study. Not all PSP were pathologically confirmed, although pathological confirmation was available in a significant subset (of the 1,718 PSP individuals, 1,441 were autopsy-confirmed and 277 were clinically-diagnosed). Additionally, the majority of control samples in this study were from ADSP and were initially collected as controls for AD studies. As ADSP is a dataset composed of multiple cohorts from diverse sources, it is imperative to ensure that any observed allele frequency differences between controls and cases can be attributed to the disease itself rather than sample selection biases arising from technical artifacts or batch effects. To mitigate the risk of false reports, we meticulously examined the allele frequencies of both cases and controls, especially in relation to novel and significant signals.

This work represents an important first step; future work is necessary to further delineate the rare genetic risk in PSP harbored in coding and noncoding regions. These results may come to fruition as additional genomic analytical methods are developed, sample size increased, and orthogonal genomic data are integrated. While PSP is rare, it is the most common primary tauopathy, and studying this disease is critical to understanding common pathological mechanisms across tauopathies. Further work to include individuals with diverse ancestry background will also improve our understanding of genetic architecture of the disease.

## Conclusion

In conclusion, this study significantly advances our understanding of the genetic basis of PSP through WGS from this study. Previous GWAS signals were validated, and *APOE2* was found to the risk allele for PSP from the analysis of common SNVs and indels. Additionally, the analysis of rare SNVs/indels and SVs has revealed additional genetic targets, including *ZNF592*, *IGH*, *PCMT1*, *CYP2A13*, and *SMCP*, opening new avenues for investigating disease mechanisms and potential therapeutic interventions.

## Declarations

### Ethics approval and consent to participate

#### Consent for publication

Not applicable.

### Availability of data and materials

NIAGADS Data Sharing Service (https://dss.niagads.org/) https://github.com/whtop/PSP-Whole-Genome-Sequencing-Analysis

### Competing interests

Laura Molina-Porcel received income from Biogen as a consultant in 2022. Gesine Respondek is now employed by Roche (Hoffmann-La Roche, Basel, Switzerland) since 2021. Her affiliation whilst completing her contribution to this manuscript was German Center for Neurodegenerative Diseases (DZNE), Munich, Germany. Thomas G Beach is a consultant for Aprinoia Therapeutics and a Scientific Advisor and stock option holder for Vivid Genomics. Huw Morris is employed by UCL. In the last 12 months he reports paid consultancy from Roche, Aprinoia, AI Therapeutics and Amylyx; lecture fees/honoraria - BMJ, Kyowa Kirin, Movement Disorders Society. Huw Morris is a co-applicant on a patent application related to C9ORF72 - Method for diagnosing a neurodegenerative disease (PCT/GB2012/052140). Giovanni Coppola is currently an employee of Regeneron Pharmaceuticals. Alison Goate serves on the SAB for Genentech and Muna Therapeutics.

## Funding

This work was supported by NIH 5UG3NS104095, the Rainwater Charitable Foundation, and CurePSP. HW and PLC are supported by RF1-AG074328, P30-AG072979, U54-AG052427 and U24-AG041689. TSC is supported by NIH K08AG065519 and the Larry L Hillblom Foundation 2021-A-005-SUP. KF was supported by CurePSP 685-2023-06-Pathway and K01 AG070326. MG is supported by P30 AG066511. BFG and KLN are supported by P30 AG072976 and R01 AG080001. TGB and GES are supported by P30AG072980. IR is supported by 2R01AG038791-06A, U01NS100610, R25NS098999, U19 AG063911-1 and 1R21NS114764-01A1. OR is support by U54 NS100693. DG is supported by P30AG062429. ALB is supported by U19AG063911, R01AG073482, R01AG038791, and R01AG071756. BLM is supported by P01 AG019724, R01 AG057234 and P0544014. VMV is supported by P01-AG-066597, P01-AG-017586. HRM is supported by CurePSP, PSPA, MRC, and Michael J Fox Foundation. RDS is supported by CurePSP, PSPA, and Reta Lila Weston Trust. JFC is supported by R01 AG054008, R01 NS095252, R01 AG060961, R01 NS086736, R01 AG062348, P30 AG066514, the Rainwater Charitable Foundation / Tau Consortium, Karen Strauss Cook Research, and Scholar Award, Stuart Katz & Dr. Jane Martin. AMG is supported by the Tau Consortium and U54-NS123746. YYL is supported by U54-AG052427; U24-AG041689. LSW is supported by U01AG032984, U54AG052427, and U24AG041689. GUH was funded by the Deutsche Forschungsgemeinschaft (DFG, German Research Foundation) under Germany’s Excellence Strategy within the framework of the Munich Cluster for Systems Neurology (EXC 2145 SyNergy – ID 390857198); Deutsche Forschungsgemeinschaft (DFG, HO2402/18-1 MSAomics); German Federal Ministry of Education and Research (BMBF, 01KU1403A EpiPD; 01EK1605A HitTau; 01DH18025 TauTherapy). DHG is supported by 3UH3NS104095, Tau Consortium. WPL is supported by RF1-AG074328; P30-AG072979; U54-AG052427; U24-AG041689. Cases from Banner Sun Health Research Institute were supported by the NIH (U24 NS072026, P30 AG19610 and P30AG072980), the Arizona Department of Health Services (contract 211002, Arizona Alzheimer’s Research Center), the Arizona Biomedical Research Commission (contracts 4001, 0011, 05-901 and 1001 to the Arizona Parkinson’s Disease Consortium) and the Michael J. Fox Foundation for Parkinson’s Research. The Mayo Clinic Brain Bank is supported through funding by NIA grants P50 AG016574, CurePSP Foundation, and support from Mayo Foundation.

## Data Availability

Data from the ADSP are available to qualified investigators via the National Institute on Aging Genetics of Alzheimer's Disease Data Storage Site (NIAGADS) (https://dss.niagads.org/).

## Acknowledgements

This project is supported by CurePSP, courtesy of a donation from the Morton and Marcine Friedman Foundation. We are indebted to the Biobanc-Hospital Clinic-FRCB-IDIBAPS and Center for Neurodegenerative Disease Research at Penn for samples and data procurement. The PSP genetics study group is a multisite collaboration including: German Center for Neurodegenerative Diseases (DZNE), Munich; Department of Neurology, LMU Hospital, Ludwig-Maximilians-Universität (LMU), Munich, Germany (Franziska Hopfner, Günter Höglinger); German Center for Neurodegenerative Diseases (DZNE), Munich; Center for Neuropathology and Prion Research, LMU Hospital, Ludwig-Maximilians-Universität (LMU), Munich, Germany (Sigrun Roeber, Jochen Herms); Justus-Liebig-Universität Gießen, Germany (Ulrich Müller); MRC Centre for Neurodegeneration Research, King’s College London, London, UK (Claire Troakes); Movement Disorders Unit, Neurology Department and Neurological Tissue Bank and Neurology Department, Hospital Clínic de Barcelona, University of Barcelona, Barcelona, Catalonia, Spain (Ellen Gelpi; Yaroslau Compta); Department of Neurology and Netherlands Brain Bank, Erasmus Medical Centre, Rotterdam, The Netherlands (John C. van Swieten); Division of Neurology, Royal University Hospital, University of Saskatchewan, Canada (Alex Rajput); Australian Brain Bank Network in collaboration with the Victorian Brain Bank Network, Australia (Fairlie Hinton), Department of Neurology, Hospital Ramón y Cajal, Madrid, Spain (Justo García de Yebenes). The acknowledgement of PSP cohorts is listed below, whereas the acknowledgement of ADSP cohorts for control samples can be found in the supplementary materials. The Genotype-Tissue Expression (GTEx) Project was supported by the Common Fund of the Office of the Director of the National Institutes of Health, and by NCI, NHGRI, NHLBI, NIDA, NIMH, and NINDS. The data used for the analyses described in this manuscript were obtained from: https://gtexportal.org/home/datasets the GTEx Portal on 1/27/2022. We also thank to Drs. Murray Grossman and Hans Kretzschmar for their valuable contribution to this work.

## AMP-AD (sa000011) data

Mayo RNAseq Study-Study data were provided by the following sources: The Mayo Clinic Alzheimer’s Disease Genetic Studies, led by Dr. Nilufer Ertekin-Taner and Dr. Steven G. Younkin, Mayo Clinic, Jacksonville, FL using samples from the Mayo Clinic Study of Aging, the Mayo Clinic Alzheimer’s Disease Research Center, and the Mayo Clinic Brain Bank. Data collection was supported through funding by NIA grants P50 AG016574, R01 AG032990, U01 AG046139, R01 AG018023, U01 AG006576, U01 AG006786, R01 AG025711, R01 AG017216, R01 AG003949, NINDS grant R01 NS080820, CurePSP Foundation, and support from Mayo Foundation. Study data includes samples collected through the Sun Health Research Institute Brain and Body Donation Program of Sun City, Arizona. The Brain and Body Donation Program is supported by the National Institute of Neurological Disorders and Stroke (U24 NS072026 National Brain and Tissue Resource for Parkinson’s Disease and Related Disorders), the National Institute on Aging (P30 AG19610 Arizona Alzheimer’s Disease Core Center), the Arizona Department of Health Services (contract 211002, Arizona Alzheimer’s Research Center), the Arizona Biomedical Research Commission (contracts 4001, 0011, 05-901 and 1001 to the Arizona Parkinson’s Disease Consortium) and the Michael J. Fox Foundation for Parkinson’s Research.

## PSP-NIH-CurePSP-Tau (sa000015) data

This project was funded by the NIH grant UG3NS104095 and supported by grants U54NS100693 and U54AG052427. Queen Square Brain Bank is supported by the Reta Lila Weston Institute for Neurological Studies and the Medical Research Council UK. The Mayo Clinic Florida had support from a Morris K. Udall Parkinson’s Disease Research Center of Excellence (NINDS P50 #NS072187), CurePSP and the Tau Consortium. The samples from the University of Pennsylvania are supported by NIA grant P01AG017586.

## PSP-CurePSP-Tau (sa000016) data

This project was funded by the Tau Consortium, Rainwater Charitable Foundation, and CurePSP. It was also supported by NINDS grant U54NS100693 and NIA grants U54NS100693 and U54AG052427. Queen Square Brain Bank is supported by the Reta Lila Weston Institute for Neurological Studies and the Medical Research Council UK. The Mayo Clinic Florida had support from a Morris K. Udall Parkinson’s Disease Research Center of Excellence (NINDS P50 #NS072187), CurePSP and the Tau Consortium. The samples from the University of Pennsylvania are supported by NIA grant P01AG017586. Tissues were received from the Victorian Brain Bank, supported by The Florey Institute of Neuroscience and Mental Health, The Alfred and the Victorian Forensic Institute of Medicine and funded in part by Parkinson’s Victoria and MND Victoria. We are grateful to the Sun Health Research Institute Brain and Body Donation Program of Sun City, Arizona for the provision of human biological materials (or specific description, e.g. brain tissue, cerebrospinal fluid). The Brain and Body Donation Program is supported by the National Institute of Neurological Disorders and Stroke (U24 NS072026 National Brain and Tissue Resource for Parkinson’s Disease and Related Disorders), the National Institute on Aging (P30 AG19610 Arizona Alzheimer’s Disease Core Center), the Arizona Department of Health Services (contract 211002, Arizona Alzheimer’s Research Center), the Arizona Biomedical Research Commission (contracts 4001, 0011, 05-901 and 1001 to the Arizona Parkinson’s Disease Consortium) and the Michael J. Fox Foundation for Parkinson’s Research. Biomaterial was provided by the Study Group DESCRIBE of theClinical Research of the German Center for Neurodegenerative Diseases (DZNE). <u>PSP_UCLA (sa000017) data</u>: Thank to the AL-108-231 investigators, Adam L Boxer, Anthony E Lang, Murray Grossman, David S Knopman, Bruce L Miller, Lon S Schneider, Rachelle S Doody, Andrew Lees, Lawrence I Golbe, David R Williams, Jean-Cristophe Corvol, Albert Ludolph, David Burn, Stefan Lorenzl, Irene Litvan, Erik D Roberson, Günter U Höglinger, Mary Koestler, Cliff ord R Jack Jr, Viviana Van Deerlin, Christopher Randolph, Iryna V Lobach, Hilary W Heuer, Illana Gozes, Lesley Parker, Steve Whitaker, Joe Hirman, Alistair J Stewart, Michael Gold, and Bruce H Morimoto.

## Authors’ contribution

Study design: TSC, DD, GUH, GDS, DHG, and WPL. Sample collection, brain biospecimens, and neuropathological examinations: TSC, CM, LM, AR, PPDD, NLB, MG, LDK, JCVS, ED, BFG, KLN, CT, JGdY, ARG, TM, WHO, GR, UM, FH, TA, SR, PP, AB, AD, ILB, TGC, GES, LNH, IL, RR, OR, DG, ALB, BLM, WWS, VMVD, EBL, CLW, HM, JH, RdS, JFC, AMG, GC, and DHG. Genotype or phenotype acquisition: HW, TSC, VP, LVB, KF, AN, LSW, GDS, DHG, and WPL. Variant detection and variant quality check: HW, TSC, VP, LVB, KF, YYL, and WPL. Statistical analyses and interpretation of results: HW, TSC, KF, AN, GDS, DHG, and WPL. Experimental validation: BAD and PLC. Draft of the manuscript: HW, TSC, GDS, DHG, and WPL. All authors read, critically revised, and approved the manuscript.

## Notes

### Summary of Updates

Evaluation of bias of sample collection for APOE impact. Add a few contributors as coauthors.

